# The relationship between voluntary and involuntary muscle contractile properties in young women

**DOI:** 10.1101/2024.10.30.24316419

**Authors:** Madison J. Fry, William S. Zoughaib, Richard L. Hoffman, Andrew R. Coggan

## Abstract

Isokinetic dynamometry and neuromuscular electrical stimulation (NMES) are two commonly used approaches for quantifying muscle contractile properties. Few studies, however, have investigated the relationship between such testing procedures, particularly in women.

**PURPOSE:** To determine the relationship between voluntary isometric and isokinetic torque and torque during involuntary, electrically evoked contractions of the knee extensor muscles.

**METHODS:** Thirty young women (age 23 ± 5 y) performed maximal knee extensions on an isokinetic dynamometer at angular velocities of 0, 1.57, 3.14, 4.71, and 6.28 rad/s. Following this testing, NMES of the quadriceps (400 V, 200 µs) was used to determine unpotentiated and potentiated twitch contractile properties. The quadriceps were also stimulated with 1 s trains at 1, 5, 10, 15, 20, 25, 30, 35, 40, 60, 80, and 100 Hz to determine the torque-frequency relationship.

**RESULTS:** Voluntary torques at 1.57 and 3.14 rad/s were significantly correlated (i.e., multiplicity-adjusted P≤0.01) with the rate of torque development during potentiated twitches (r = 0.60 and 0.55, respectively). No other significant correlations were found between voluntary and involuntary muscle contractile properties, including various measures of the torque-frequency relationship.

**CONCLUSION:** Although there is some relationship between voluntary and NMES indices of muscle contractility, such results are only moderately well-correlated at best. The two techniques should therefore be considered complementary rather than interchangeable.

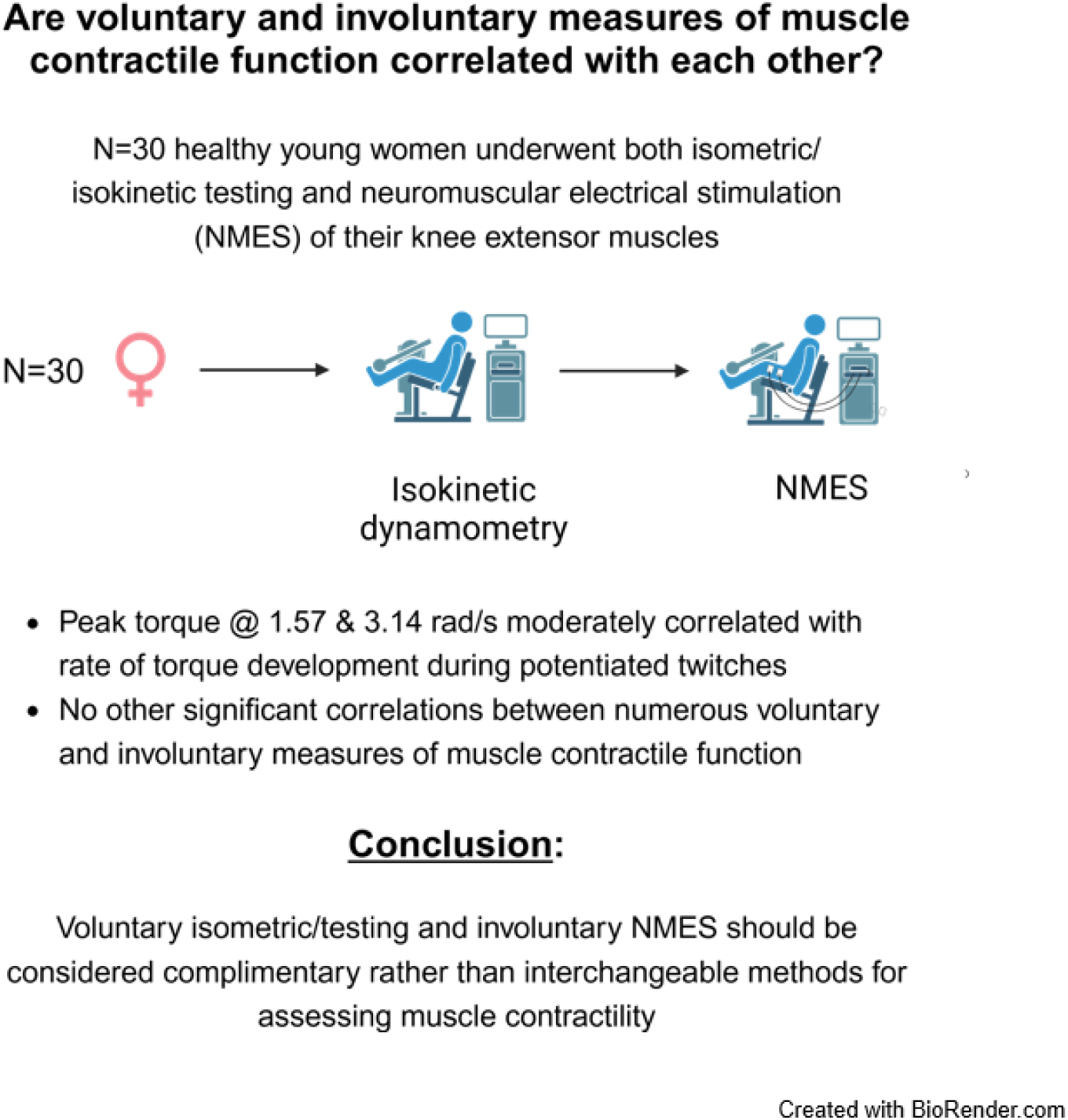

## Introduction

The contractile properties of skeletal muscle play a crucial role in determining athletic performance (Baker et al. 2001; Sleivert and Taingaue 2004). They are also an important determinant of the ability to perform ordinary activities of daily living (e.g., rising from a chair) and hence the quality of life, independence, and risk of falls/fractures in individuals with compromised muscle function (e.g., older persons) (Pojednic et al. 2012; Reid and Fielding 2012). It is therefore important to understand the advantages, limitations, and interrelationships between different methods of quantifying muscle contractility in both healthy and clinical populations, as this may influence the development of exercise training methods, nutritional interventions, and/or pharmacological therapies aimed at enhancing muscular performance.

Muscle contractile function can be assessed in various ways. Most often, studies have focused upon evaluating strength, i.e., the maximal force generating capacity of muscle, either by assessing maximal isometric force production (e.g., during a handgrip) or by measuring (or estimating) the maximum weight that can be lifted a single time against gravity (i.e., an individual’s one repetition-maximum (1 RM)) during a particular movement (e.g., Roschel et al. 2021). However, maximal muscle shortening velocity, and hence maximal power (i.e., the product of force and velocity), is a better predictor of both athletic performance and ordinary physical function (e.g., ability to climb stairs) than strength (Baker et al. 2001; Pojednic et al. 2012; Reid and Fielding 2012; Sleivert and Taingaue 2004). Thus, since the 1970s isokinetic dynamometry has been routinely used to quantify the force (torque) a muscle or muscle group can produce while controlling for velocity. Although not without its limitations (e.g., torque artifacts, inability to reach the target velocity (Lanza et al. 2003; Osternig 1986; Sapega et al. 1982)), isokinetic dynamometry is widely considered the clinical gold standard for assessing voluntary muscle function (Baltzopoulos et al. 2012; Osternig 1986).

Another approach for determining muscle contractility is to elicit involuntary contractions via neuromuscular electrical stimulation (NMES), i.e., via application of electrical stimuli to motor nerves or to the muscle itself. NMES enables determination of intrinsic muscle properties, e.g., twitch characteristics, force-frequency relationship, that cannot be assessed during voluntary contractions and that may provide mechanistic insight into the determinants of muscle function. NMES measures are also independent of an individual’s volitional effort and therefore tend to be less variable than voluntary measures (Clark et al. 2007; Jenkins et al. 2014). On the other hand, NMES results in non-selective recruitment of both fast and slow muscle fibers/motor units, versus the orderly, slow-to-fast hierarchy that exists during voluntary movements due to Henneman’s size (Bickel et al. 2011). Results obtained using NMES may therefore reflect sampling from a faster motor unit pool than that recruited during even maximal voluntary contractions. Finally, another disadvantage of NMES is that shocking the motor nerve or muscle directly can be painful (Neyroud et al. 2015), especially with higher currents and/or at higher frequencies as is required to maximize muscle recruitment (Kendall et al. 2006).

Despite the widespread use of both isometric/isokinetic testing and NMES, the exact relationship between these two approaches for assessing muscle contractile function is not entirely clear. Results obtained using the two techniques have generally paralleled each other in some situations (e.g., when comparing men vs. women (Behm and Sale 1994), with resistance training/detraining of younger individuals (Andersen et al. 2005; Ishida et al. 1990), when studying fatigue (e.g., Babault et al. 2006; Michaut et al. 2003)), but not others (e.g., passive (Trajano et al. 2013) or dynamic (Skurvydas et al. 2007) stretching, with resistance training of older adults (Clark et al. 2021)). Similarly, while some have observed moderately high correlations between selected voluntary and involuntary measures of muscle function within a given individual (e.g., Wrucke et al. 2024), others have not (e.g., Phillips et al. 2022). In part, these findings likely reflect the varying contribution of neural factors during voluntary but not involuntary testing. Nonetheless, a comprehensive assessment of the relationship between the results of isometric/isokinetic testing and NMES measurements of muscle contractile properties appears to be lacking.

The aim of this study was to investigate the interrelationships between various commonly reported measures obtained via isometric/isokinetic testing and NMES, respectively, in young, healthy, normally active women. We studied women because they have not been the focus of prior research and because sex-related differences in fiber type size/distribution (Nuzzo 2024) and adipose tissue layer thickness (Doheny et al. 2008; Maffiuletti et al. 2008; Medeiros et al. 2015; Miller et al. 2008) could potentially influence how well the two methods might relate to each other.

## Methods

### Participants

Potential participants were recruited by word of mouth and flyers placed around the university campus. Exclusion criteria were age <18 or >44 y; irregular menstrual cycle (average length <21 or >35 d); having missed more than three consecutive periods in the last 12 mo; currently pregnant; currently taking hormonal contraceptives or on hormone replacement therapy; currently using antibiotics; current smoker; diagnosis of epilepsy or presence of pacemaker, other implantable cardiac device, or copper IUD (safety procedures for NMES); resting blood pressure >140/90 mmHg; an answer of yes to any of the seven general health questions of the Physical Activity Readiness Questionnaire (PAR-Q); an International Physical Activity Questionnaire (IPAQ) score >3000 met-min/wk; or the inability to provide informed consent. After screening 50 participants, 32 were determined to be eligible, of whom 30 ultimately completed the study. (One participant became lightheaded during the NMES protocol and the experiment was stopped, whereas voluntary muscle function data from another were lost due to a computer malfunction.) Participant characteristics are presented in Table 1. The Human Subjects Office at Indiana University approved this study and written informed consent was obtained from each individual.

**Table 1.** Participant characteristics.

|  |  |
| --- | --- |
| Age (y) | 23 ± 5 |
| Height (m) | 1.64 ± 0.07 |
| Mass (kg) | 64.7 ± 11.2 |
| BMI (kg/m<sup>2</sup>) | 24.1 ± 4.0 |
| IPAQ Score (met-min/wk) | 1344 ± 769 |

IPAQ, International Physical Activity Questionnaire. Values are mean ± SD for n=30.

### Experimental procedures

Participants reported to the Exercise Physiology Laboratory at Indiana University Indianapolis. After completion of the informed consent process and PAR-Q, IPAQ, and medical history questionnaires to determine eligibility, their height, weight, and seated heart rate and blood pressure were measured. An isokinetic dynamometer (Biodex System 4 Pro, Biodex Medical Systems, Shirley, NY) was then used to determine the maximal voluntary isometric and isokinetic torque of the knee extensor muscles of the participant’s dominant leg, as previously described in detail (Coggan et al. 2015). Briefly, after adjustment of the dynamometer and placement of straps over the participant’s waist, torso, and thigh to restrict extraneous movement, they were asked to perform three maximal 5 s isometric contractions, with 15 s rest in between. Following 2 min of rest, they performed three maximal knee extensions at angular velocities of 1.57, 3.14, 4.71, and 6.28 rad/s (90, 180, 270, and 360 °/s), in that order, with 2 min of rest between each set. Participants were instructed to extend their knee as “hard and fast as possible” (Rendos et al. 2015) during all contractions and strong verbal encouragement was provided throughout the testing.

Following completion of the above testing, NMES was used to determine the involuntary contractile properties of the quadriceps muscle. The participant’s thigh was first shaved and abraded, to minimize impedance of the skin, after which two 5.08 cm x 8.89 cm self-adhesive electrodes (Dura-Stick Plus, Chattanooga Medical Supply, Chattanooga, TN) were applied to the leg. The cathode was placed longitudinally over the vastus lateralis, 3-5 cm distal to the inguinal crease, whereas the anode was placed longitudinally over the vastus medialis, 3-5 cm proximal to the border of the patella (Brooks et al. 1990). A constant current stimulator (Digitimer DS-7AH, Hertfordshire, UK) was then used to elicit isometric contractions at a knee joint angle of 90°, with torque measured at 1000 Hz using the isokinetic dynamometer interfaced with a computer via a 16 channel data acquisition system (Biopac MP160, Biopac, Goleta, CA). First, single stimuli (400 V, 200 µ) of increasing current were applied at 3-5 s intervals until a plateau or decrease in twitch torque was observed. After a brief rest, the amperage resulting in the highest torque (i.e., 294±79 mA; range 175- 475 mA) was then used to elicit four unpotentiated twitches at approximately 1 s intervals. The participant then performed a 6 s maximal voluntary contraction (MVC), immediately after which four potentiated twitches were elicited, also at approximately 1 s intervals. A single stimulus was also applied during the MVC to determine voluntary activation using the interpolated twitch technique (ITT), as described in greater detail below. This sequence was repeated twice, with at least 10 min in between to allow for reversal of the potentiation. After a further 10 min of rest, participants were stimulated at 100 Hz with 1 s trains of increasing current to determine the amperage eliciting approximately 33% of the maximum torque observed during the preceding MVCs. This amperage (i.e., 73±16 mA; range 45-100 mA) was then applied in 1 s trains at 3-5 s intervals at (in order) 1, 5, 10, 15, 20, 25, 30, 35, 40, 60, 80, and 100 Hz to determine the torque-frequency relationship. (Note that the shape of this relationship is independent of the elicited torque between 25 and 50% of MVC, and only changes slightly even at 80% of MVC (Binder-Macleod et al. 1995)).

### Data analysis

Torque data collected during the voluntary isometric and isokinetic testing were windowed and filtered using Biodex Advantage BX version 5.3.06, then manually checked to verify the absence of artifacts. The highest torque generated at each velocity was then used to calculate peak power at that velocity, with the resulting peak power versus velocity relationship fit with an inverted parabola (average r = 0.993) to determine the maximal power (P_max_) and maximal velocity (V_max_) of the knee extensors, as previously described (Coggan et al. 2015).

Torque data collected during NMES were analyzed using Biopac AcqKnowledge version 5.08. Data were first smoothed using a 10 ms rolling average filter. Peak twitch torque (PTT), time to peak torque (TPT), and one-half relaxation time (HRT) were then manually extracted for each twitch. Maximal rates of torque development (RTD) and relaxation (RR) were also determined based on the first derivative of the smoothed torque signal over a 20 ms interval. The eight values obtained for each of these parameters under each condition (i.e., unpotentiated and potentiated) were then averaged and these averages used in all subsequent calculations.

Voluntary activation (VA) during the MVCs was determined by measuring the smoothed torque immediately before (*torque pre*) the single stimulus applied during the MVC and highest value recorded during the next 150 ms (*torque post*). VA was calculated as:

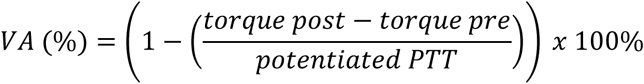

The highest of the two values was taken as most representative of the participant’s maximal volitional effort.

After smoothing, the peak torque during each of the 1 s trains at 1-100 Hz was manually extracted and expressed relative to the peak torque measured at 100 Hz. The data were then fit to a four-parameter sigmoidal function (average r = 0.989) to determine the frequency resulting in 50% of the predicted plateau in torque (F_50_) and the steepness of the curve at this point (Hill n) (Mela et al. 2001, Russ et al. 2009):

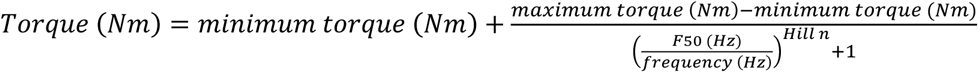

The twitch:tetanus ratio was then calculated by dividing the fitted minimum torque by the fitted maximum torque.

### Statistical analysis

Statistical analyses were performed using GraphPad Prism version 10.2.2 (GraphPad Software, La Jolla, CA). Interrelationships between various measures of muscle contractile function during voluntary testing and NMES were determined via a correlation matrix. The two-stage linear step procedure of Benjamini, Krieger, and Yekulietli (Benjamini et al. 2006) was used for multiplicity correction, with the false discovery rate (FDR) set to 1%. After this correction, any P value <0.00173, corresponding to an r of > | 0.540 |, was considered statistically significant.

## Results

Results from the voluntary isometric/isokinetic testing are shown in Table 2, whereas the twitch and torque-frequency data obtained via NMES are shown in Table 3. The correlation matrix is shown in Fig. 1. For all parameters, the highest and lowest values varied by a factor of at least ∼2, minimizing the risk of any restriction-in-range errors.

**Fig. 1.**
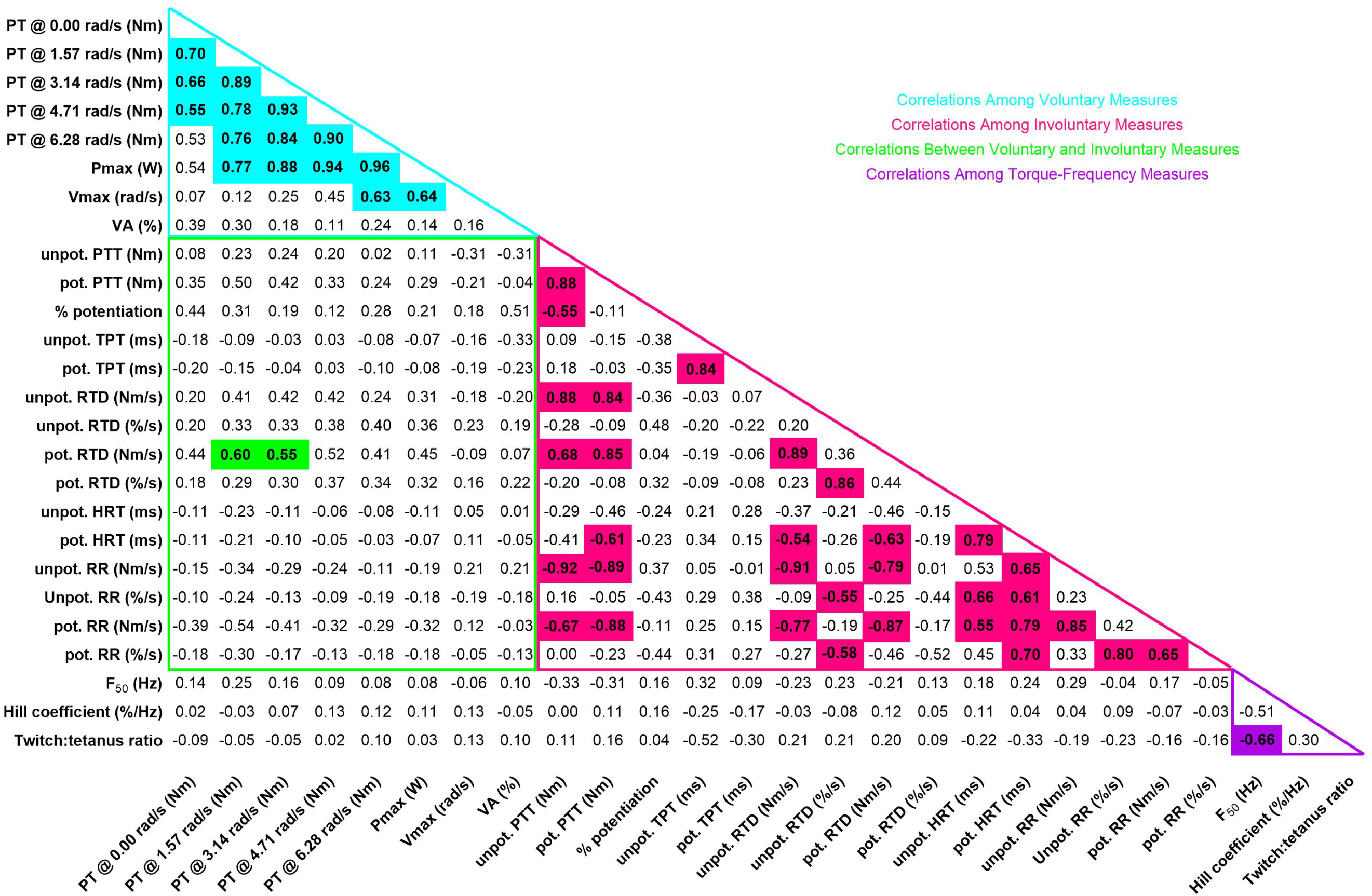
Correlations between voluntary and involuntary measures of muscle contractile function. Multiplicity-adjusted P values that are statistically significant are shown in bold on colored background. See text for details

**Table 2.** Muscle contractile properties determined during voluntary exercise.

| <u>Parameter</u> | <u>Mean ± SD</u> | <u>Range</u> |
| --- | --- | --- |
| Peak torque @ 0.00 rad/s (Nm) | 141.8 ± 29.9 | 65.1 to 193.7 |
| Peak torque @ 1.57 rad/s (Nm) | 108.1 ± 29.7 | 43.5 to 163.8 |
| Peak torque @ 3.14 rad/s (Nm) | 84.7 ± 24.5 | 27.7 to 137.9 |
| Peak torque @ 4.71 rad/s (Nm) | 69.2 ± 21.2 | 19.3 to 112.3 |
| Peak torque @ 6.28 rad/s (Nm) | 55.3 ± 19.8 | 8.7 to 90.8 |
| Pmax (W) | 361 ± 117 | 105 to 603 |
| Vmax (rad/s) | 12.2 ± 3.3 | 7.4 to 18.6 |
| Voluntary activation (%) | 86.0 ± 11.2 | 45.8 to 99.0 |

Pmax, maximal knee extensor power. Vmax, maximal velocity of knee extension. Values are
mean ± SD for n=30.

**Table 3.**
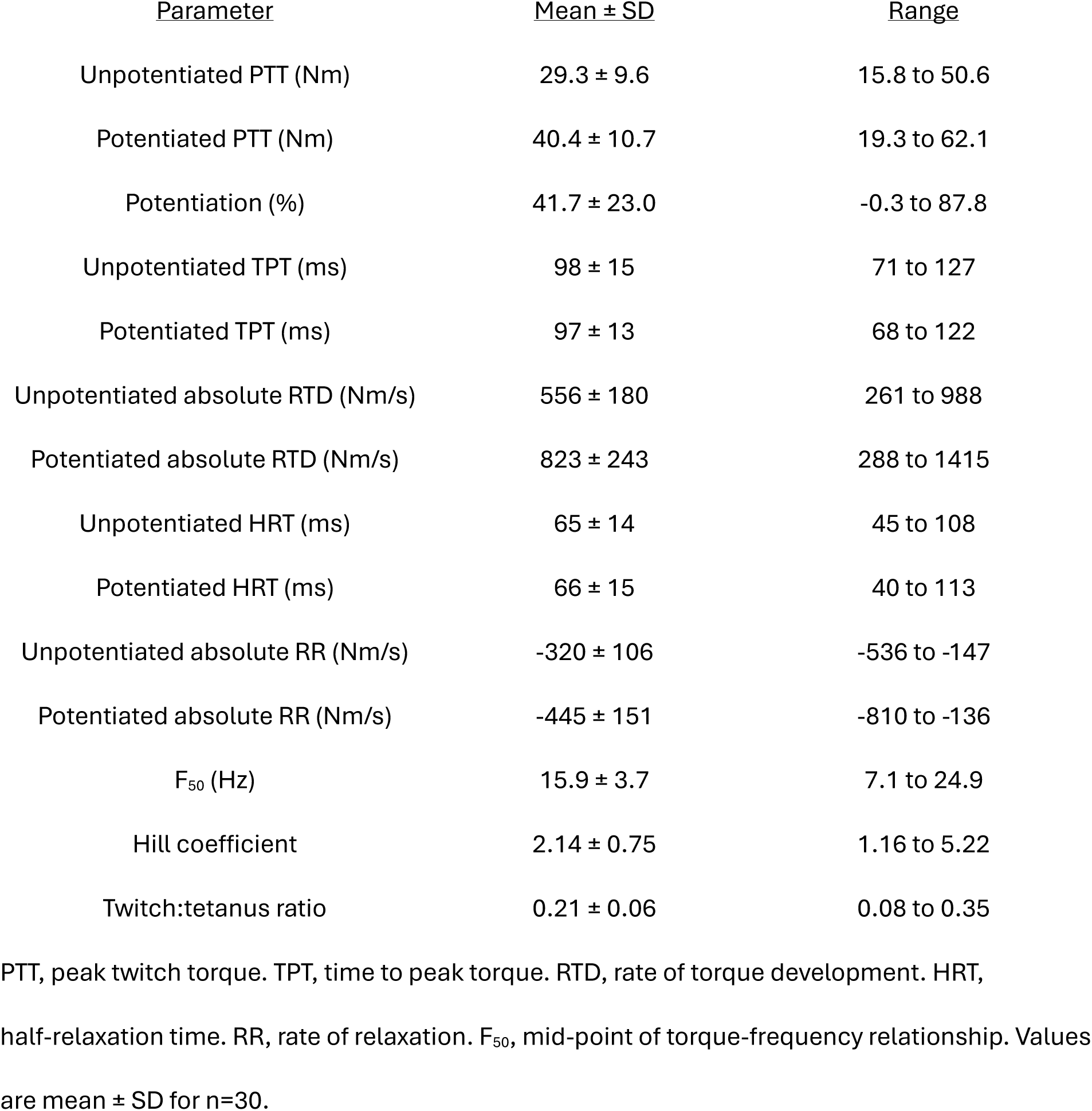
Muscle contractile properties determined during involuntary exercise.

| <u>Parameter</u> | <u>Mean ± SD</u> | <u>Range</u> |
| --- | --- | --- |
| Unpotentiated PTT (Nm) | 29.3 ± 9.6 | 15.8 to 50.6 |
| Potentiated PTT (Nm) | 40.4 ± 10.7 | 19.3 to 62.1 |
| Potentiation (%) | 41.7 ± 23.0 | -0.3 to 87.8 |
| Unpotentiated TPT (ms) | 98 ± 15 | 71 to 127 |
| Potentiated TPT (ms) | 97 ± 13 | 68 to 122 |
| Unpotentiated absolute RTD (Nm/s) | 556 ± 180 | 261 to 988 |
| Potentiated absolute RTD (Nm/s) | 823 ± 243 | 288 to 1415 |
| Unpotentiated HRT (ms) | 65 ± 14 | 45 to 108 |
| Potentiated HRT (ms) | 66 ± 15 | 40 to 113 |
| Unpotentiated absolute RR (Nm/s) | -320 ± 106 | -536 to -147 |
| Potentiated absolute RR (Nm/s) | -445 ± 151 | -810 to -136 |
| F<sub>50</sub> (Hz) | 15.9 ± 3.7 | 7.1 to 24.9 |
| Hill coefficient | 2.14 ± 0.75 | 1.16 to 5.22 |
| Twitch:tetanus ratio | 0.21 ± 0.06 | 0.08 to 0.35 |

PTT, peak twitch torque. TPT, time to peak torque. RTD, rate of torque development. HRT, half-relaxation time. RR, rate of relaxation. F<sub>50</sub>, mid-point of torque-frequency relationship. Values are mean ± SD for n=30.

*Correlations among voluntary measures:*

Isometric peak torque (i.e., at 0.00 rad/s) was significantly correlated with isokinetic peak torque at 1.57-4.71 rad/s (i.e., r = 0.553-0.701, multiplicity-adjusted P = 0.00920-0.000154), but not at 6.28 rad/s. Peak torques at 1.57-6.28 rad/s were also all significantly correlated with each other (i.e., r = 0.761-0.927, p = 1.08 x 10^-5^-1.90 x 10^-11^). Correspondingly, P_max_ was significantly correlated with peak torque at 1.57-6.28 rad/s (i.e., r = 0.765- 0.958, P = 8.70 x 10^-6^-2.82 x 10^-14^), but not with isometric peak torque (Fig. 2). V_max_ was also significantly correlated with peak torque at 6.28 rad/s (i.e., r = 0.634, P = 0.00122) and with P_max_ (i.e., r = 0.640, P = 0.00104). VA did not correlate with any other measure of voluntary muscle function.

**Fig. 2.**
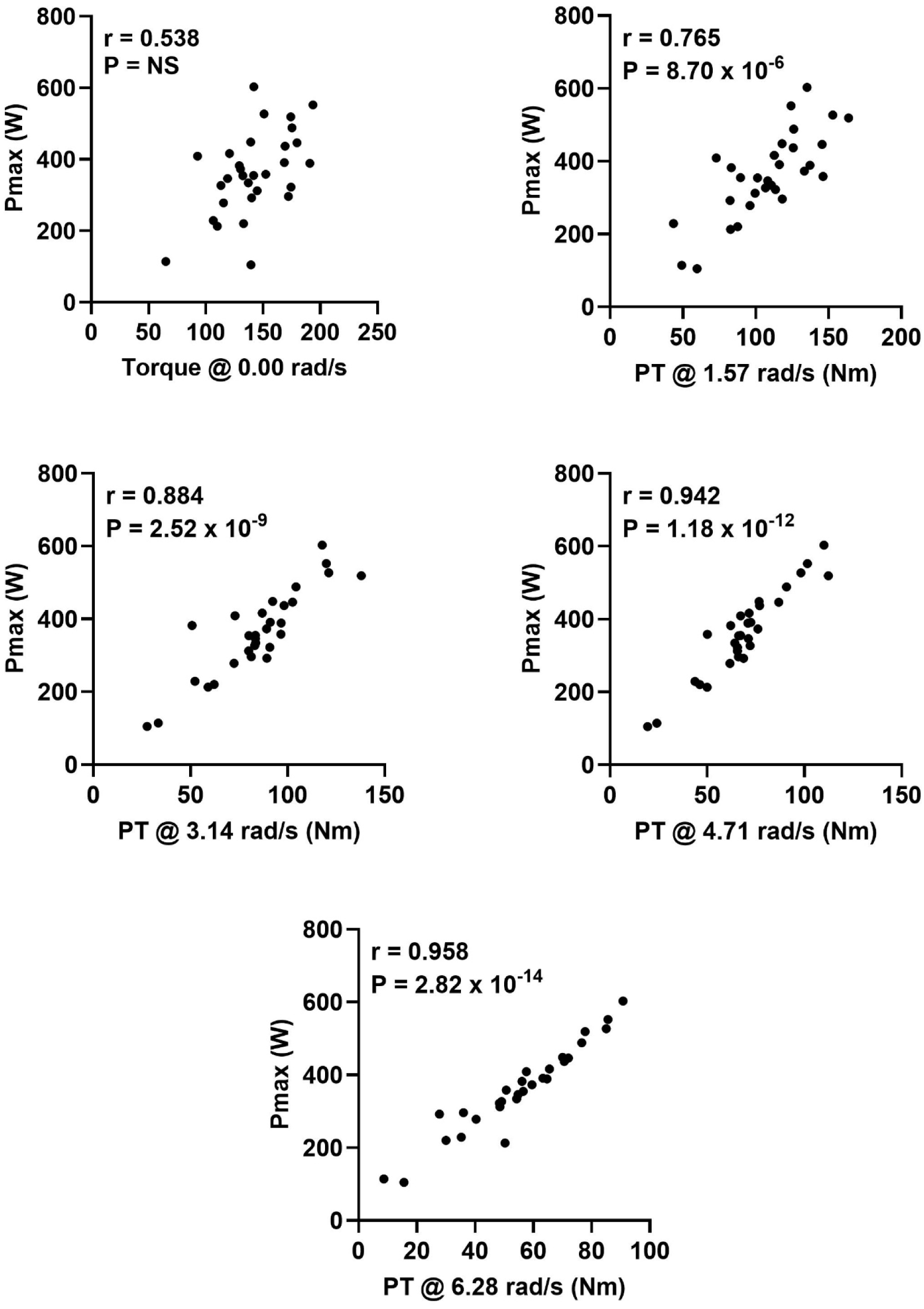
Relationship of maximal knee extensor power (Pmax) to isometric and isokinetic peak torques (PT).

### Correlations among involuntary measures

Numerous significant correlations were observed among NMES parameters, including between unpotentiated and potentiated PTT; unpotentiated and potentiated TPT; unpotentiated and potentiated absolute RTD; unpotentiated and potentiated HRT; and unpotentiated and potentiated absolute RR (i.e., r = 0.785-0.885, P = 3.30 x 10^-6^-2.52 x 10^-9^) (Fig. 3). Absolute RTD was also significantly correlated with absolute PTT in both the unpotentiated (i.e., r = 0.876, P = 5.17 x 10^-9^) and potentiated (i.e., r = 0.849, P = 5.25 x 10^-8^) states, as were the corresponding absolute RR (i.e., r = -0.916, P = 9.58 x 10^-11^ and r = -0.877, P = 5.11 x 10^-9^), respectively. On the other hand, the results of the torque-frequency testing did not correlate with any twitch parameter, although F_50_ was inversely related to the twitch-tetanus ratio (i.e., r = -0.662; P = 0.000541). Other significant correlations between NMES measures are shown in Fig. 1.

**Fig. 3.**
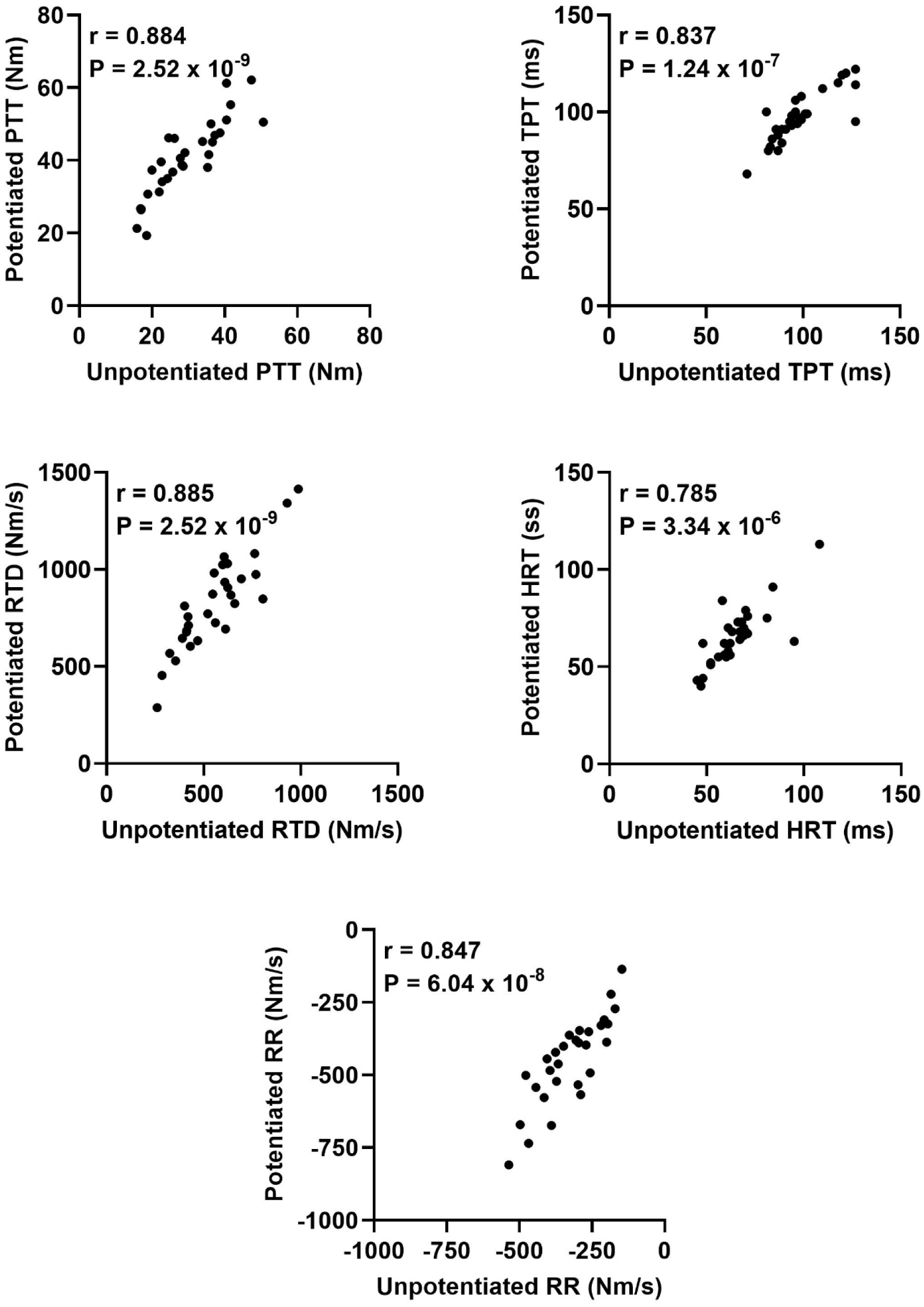
Relationship of potentiated to unpotentiated twitch characteristics.

### Correlations between voluntary and involuntary measures

Despite the internal consistency of the isometric/isokinetic and NMES measurements, as demonstrated by the numerous robust relationships within each group, only two of the 144 correlations between them were statistically significant. Specifically, peak torques at 1.57 and 3.14 rad/s were correlated with the potentiated absolute RTD (i.e., r = 0.597, P = 0.00331 and r = 0.551, P = 0.00920, respectively) (Fig. 4). No other significant correlations were observed between voluntary and involuntary measures of muscle contractile function.

**Fig. 4.**
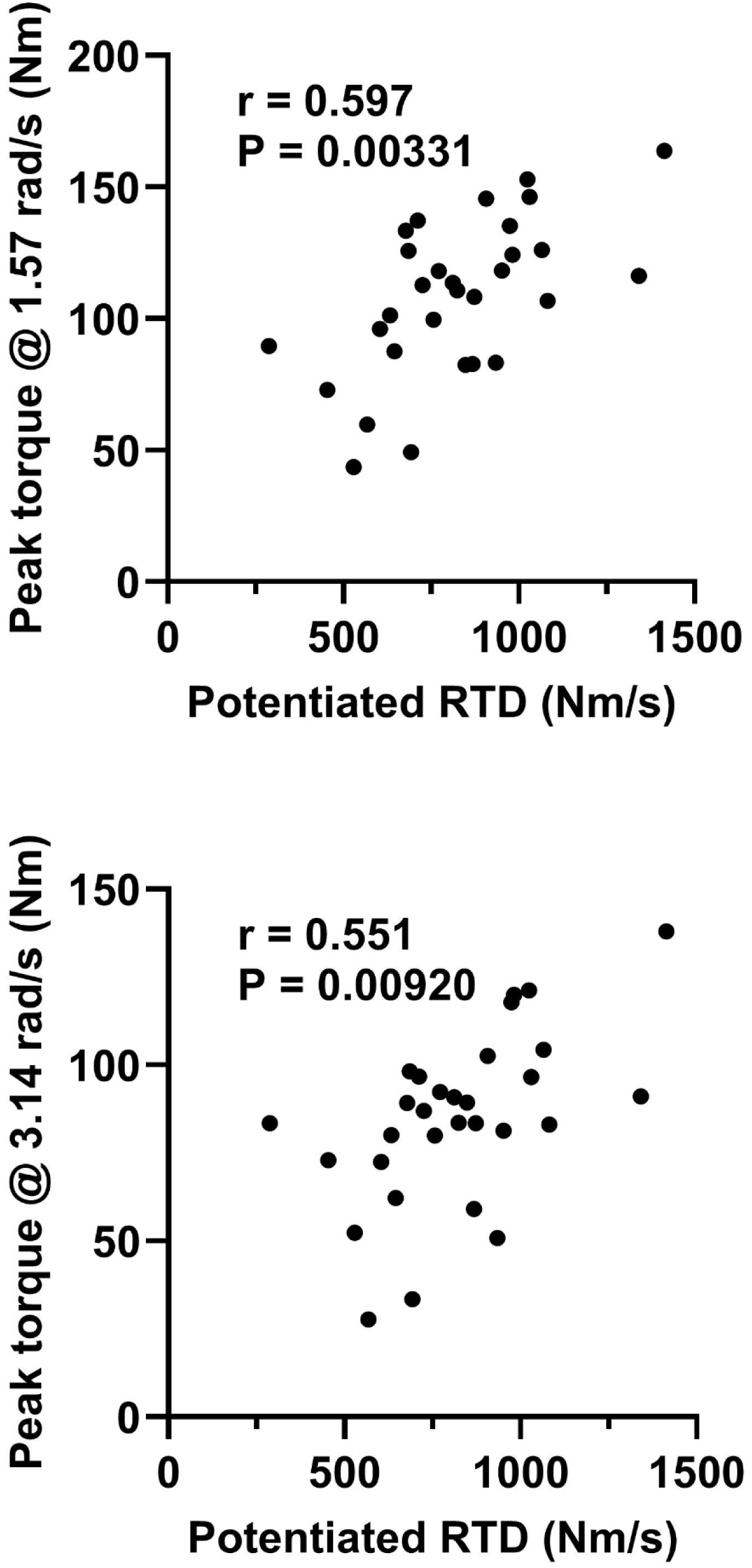
Relationship of isokinetic peak torque at 1.57 and 3.14 rad/s to the rate of torque development (RTD) during potentiated twitches.

## Discussion

Both isometric/isokinetic testing and NMES are frequently used to quantify muscle contractile function, but how well the two methods relate to each other is not entirely clear. The primary purpose of this study was therefore to determine the relationship, if any, between such measures of muscle contractility in young women. We found numerous significant correlations among various parameters obtained using either technique, but only limited correlation between the two methods. This is the first comprehensive comparison of muscle function during voluntary isometric/isokinetic vs. involuntary, i.e., electrically evoked, isometric contractions in this, or to our knowledge any, population.

An important advantage of NMES is that by stimulating the motor nerve or muscle directly, the influence of the CNS is bypassed, theoretically allowing more direct assessment of muscle contractile properties, independently from participant motivation. It is therefore tempting to conclude that the general lack of correlation observed between the various measures of voluntary and involuntary muscle contractile function are the result of varying central motor drive between participants during the former testing. However, VA as assessed using the ITT was generally high and was not significantly correlated with any other parameter, suggesting that other factors probably also played a role. These include possible differences between participants in muscle fiber type size and/or distribution, which could have influenced the extent to which NMES and voluntary contractions recruited the same motor units (Bickel et al. 2011). Similarly, differences between participants in adipose tissue layer thickness (Doheny et al. 2008; Maffiuletti et al. 2008; Medeiros et al. 2015; Miller et al. 2008) and electrode placement (Brooks et al. 1990; Pietrosimone et al. 2011; Viera et al. 2016) likely impacted the extent to which it was possible to activate the quadriceps via NMES. Finally, the twitch response of a muscle is a complex phenomenon, the size and speed of which reflects not only the rates and/or magnitudes of Ca^2+^ release, crossbridge cycling, and Ca^2+^ reuptake, but also the stiffness of the series elastic component (Josephson 1999; Rome and Lindstedt 1997). The relative importance of these factors in determining muscle force output will almost certainly differ in response to a single electrical stimulus vs. repetitive depolarization of the alpha motor neuron, as normally occurs during voluntary muscle activity. In light of the above, the fact that only low-to-moderate correlations were observed between NMES measures of muscle contractile properties and *in vivo* function is perhaps not unexpected.

As indicated previously, relatively few previous studies seem to have directly examined the relationship between results obtained using isometric/isokinetic testing vs. NMES, with the majority focusing upon the role of intrinsic muscle contractile properties as a determinant of the voluntary RTD (which we did not quantify) (e.g., Andersen and Aagaard 2006; Phillips et al. 2022). In seeming contrast to our results, however, Wrucke et al. (Wrucke et al. 2024) recently reported moderately high correlations (i.e., r = 0.758-0.827, P<0.01) between isokinetic peak power and PTT or RTD determined via NMES. Lower but still often significant correlations were also observed between isokinetic peak power and HRT. These results, however, were driven in part by pooling data from a large number of young and older men and women – in the 15 young women that they studied, the correlation between isokinetic peak power and RTD during potentiated twitches was lower (i.e., r = 0.541, unadjusted P = 0.0372) (data extracted from Fig. 7B using PlotDigitizer (plotdigitizer.com)). This is not dissimilar to the r = 0.446 (unadjusted P = 0.0134) that we observed. Thus, rather than being contradictory our data are in fact in keeping with those of Wrucke et al. (Wrucke et al. 2024).

Although we found only limited correlation between the results of isometric/isokinetic testing and NMES, there were much stronger associations among parameters obtained with one technique or the other. For example, isometric peak torque was significantly correlated with isokinetic peak torque measured at 1.57-4.71 rad/s, and peak torques at 1.57-6.28 rad/s were all also moderately-to-highly correlated with one another. However, the relationship between isometric peak torque and torque 6.28 rad/s was not significant. Consequently, there was no relationship between isometric peak torque and Pmax. These results are generally consistent with those of Knapik and Ramos (Knapik and Ramos 1980), who found in 352 young men that although knee extensor isometric peak torque was significantly correlated with isokinetic peak torque at 0.52-3.14 rad/s, the closeness of this association decreased with increasing angular velocity. More importantly, however, these results emphasize the fact that the determinants of muscular strength and power are, to a considerable degree, independent of one another (Fitts et al. 1991; Josephson 1999; Sargeant 2007). Since power is more important than strength in determining functional outcomes (Baker et al. 2001; Pojednic et al. 2012; Reid and Fielding 2012; Sleivert and Taingaue 2004), clinical studies of, e.g., the effects of nutritional supplements on muscle contractility in frail individuals should be focused on determining changes in power, instead of simply measuring changes in strength (Roschel et al. 2021).

Similar to the above, many of the parameters determined via NMES were significantly correlated with each other. Despite differences in potentiation between individuals, many of these associations were between the same twitch characteristic measured in both the unpotentiated and the potentiated states. Thus, unless specifically studying post-activation potentiation and/or its underlying mechanisms (e.g., phosphorylation of the myosin regulatory light chain (Vandenboom 2016)), there seems little reason to favor one vs. the other. Potentiated twitch parameters tended to be slightly more reproducible (data not shown), however, and therefore may provide a more sensitive indicator in response to small changes. There were also significant correlations between PTT and absolute RTD and RR during both unpotentiated and potentiated twitches, thus supporting the usual practice of normalizing RTD and RR to the size of the twitch (e.g., Ishida et al. 1990). Finally, although correlated with each other none of the parameters obtained via torque-frequency testing (i.e., F_50_, Hill coefficient, twitch:tetanus ratio) were significantly associated with any of the twitch characteristics. These findings speak to the uniqueness of the data provided by each type (i.e., twitch vs. tetanic) of NMES testing.

There are limitations to the current study. We measured numerous outcomes in a relatively small number of individuals; thus, despite correction for multiplicity via application of a stringent FDR it is possible that some of the correlations that we observed are spurious. Conversely, it is also possible that despite the wide range in both voluntary and involuntary measures some of the weaker relationships would have proved significant had we studied more participants. We also studied only the quadriceps of young, healthy, normally active women, and our results may not apply to other muscles/muscle groups or other populations (including men). Finally, our participants did not practice the isometric/isokinetic testing before data were collected, which may have resulted in extraneous variability due to lack of familiarity with the procedures. However, VA was >75% in all but two of the women, with the overall average being comparable to that reported by others using the interpolated twitch technique (e.g., Grindstaff and Threlkeld 2014; Michaut et al. 2003; Pietrosimone et al. 2011; Paris and Rice 2024; Sigrist et al. 2024). Furthermore, the power-velocity relationship determined via the isokinetic testing was well-described by the fitted curve in each individual. Together, these data suggest that we adequately assessed maximal muscle contractile properties during voluntary contractions.

In summary, we have determined the interrelationships between commonly reported indices of muscle contractile function assessed during voluntary vs. involuntary contractions, i.e., via isometric/isokinetic testing and NMES, respectively. Although internally consistent, as evidenced by the numerous correlations observed between parameters measured using one method or the other, there was very limited correlation between them. The two techniques should therefore be considered complimentary rather than interchangeable.

## Data Availability

All data produced in the present study are available upon reasonable request to the authors

## Author Contributions

MJF and ARC conceived and designed the research and wrote the manuscript. RLH prepared regulatory documents. All authors conducted experiments and read and approved the final manuscript.

## Acknowledgements

Madison J. Fry was supported by an Undergraduate Research Opportunity Program grant from the Center for Teaching and Learning at Indiana University Indianapolis.

## Abbreviations

F_50_: Mid-point of torque-frequency relationship.
HRT: Half-relaxation time
IPAQ: International Physical Activity Questionnaire
ITT: Interpolated twitch technique
MVC: Maximal voluntary contraction
NMES: Neuromuscular electrical stimulation
P_max_: Maximal knee extensor power
PAR-Q: Physical Activity Readiness Questionnaire
PT: Peak torque
PTT: Peak twitch torque
RR: Rate of relaxation
RTD: Rate of torque development
TPT: Time to peak torque
VA: Voluntary activation
V_max_: Maximal knee extensor velocity

